# Sensitivity assessment of SARS-CoV-2 PCR assays developed by WHO referral laboratories

**DOI:** 10.1101/2020.05.03.20072207

**Authors:** Sibyle Etievant, Antonin Bal, Vanessa Escuret, Karen Brengel–Pesce, Maude Bouscambert, Valérie Cheynet, Laurence Generenaz, Guy Oriol, Gregory Destras, Geneviève Billaud, Laurence Josset, Emilie Frobert, Florence Morfin, Alexandre Gaymard

**Author notes:** equal contributions (Sibyle Etievant, Antonin Bal).

## Abstract

The sensitivity of SARS-CoV-2 RT-PCR tests developed by Charité (Germany), HKU (Hong-Kong), China CDC (China), US CDC (United-States), and Institut Pasteur, Paris (France) was assessed on SARS-CoV-2 cell culture supernatants and clinical samples. Although all RT-PCR assays performed well for SARS-CoV-2 detection, RdRp Institut Pasteur (IP2, IP4), N China CDC, and N1 US CDC were found to be the most sensitive.

---

A new human coronavirus called severe acute respiratory syndrome coronavirus 2 (SARS-CoV-2) emerged in China, in December 2019 [1]. SARS-CoV-2 is responsible for the coronavirus disease 2019 (COVID-19) which was declared a pandemic on 12 March 2020 by the World Health Organization (WHO) [2]. As of 16 April, 1 991 562 cases have been reported including 130 885 deaths [3]. A sensitive diagnostic assay is crucial to limit SARS-CoV-2 spreading as it allows to early detect new cases which lead to patient isolation and contact tracing. The first SARS-CoV-2 genome was published on 10 January 2020 [4] enabling the rapid design of a real-time reverse-transcriptase polymerase chain reaction (RT-PCR) assay by Charité (Germany) [5,6]. This test was the first to be dispatched by WHO [7] and was widely implemented in clinical virology laboratories worldwide [8]. Since then, WHO has published [9] other approaches developed by referral laboratories including HKU (Hong-Kong) [10,11], China CDC (China) [12], US CDC (United-States) [13] and Institut Pasteur, Paris (France) [14]. These assays target two or three different SARS-CoV-2 gene regions, including RdRp (RNA-dependent RNA polymerase), N (nucleocapsid protein), E (envelope protein), ORF1ab nsp10 (non-structural protein 10), and ORF1b nsp14 (non-structural protein 14). In the present study, we aimed to compare the sensitivity of these different RT-PCR assays.

## Study design

Different RNA concentrations obtained by a nine-fold serial dilution of SARS-CoV-2 cell culture supernatants as well as clinical samples were first tested using each RT-PCR assay. Limit of detection (LoD) for the three most sensitive assays was then assessed. Clinical samples with low viral concentration or tested negative were further tested using these three assays.

## Coronavirus cell culture supernatant and clinical samples

Thirty-two clinical samples (nasopharyngeal aspirates) were provided by the Hospices Civils de Lyon – University Hospital, France. Eight clinical samples were tested using all PCR assays, and twenty four samples were tested only with the most sensitive assays. Samples were frozen at –80°C before extraction. A positive sample from a patient was cultivated on buffalo green monkey cells in a biosafety level 3 laboratory to collect cell culture supernatants containing SARS-CoV-2. The SARSCoV-2 culture had an infectious titer of 8.27 log_10_ TCID_50_ as assessed by the Reed and Muench statistical method [15].

## Extraction and RT-PCR

RNA extraction was performed using the EMAG^®^ platform (Biomerieux, Marcy-l’Étoile, France), according to manufacturer’s instructions. RT-PCR were performed following published instructions [5,6,10–14] which are summarised in table 1 and 2. Since the China CDC protocol does not specify polymerase, thermocycler, volume of RNA extract, and amplification cycles, the same instructions as the HKU assay were applied. RdRp IP2 and IP4 assays from Institut Pasteur, Paris (France) can be multiplexed or used in simplex [14]. Preliminary comparison on SARS-CoV-2 cell culture supernatants found that RdRp IP4 performed better when used in multiplex whereas IP2 was not significantly impacted (supplementary table 1). Since not all thermocyclers were available to us, the CFX 96 Touch™ Real-Time PCR (Biorad) was used for all RT-PCR assays.

**Table 1:**
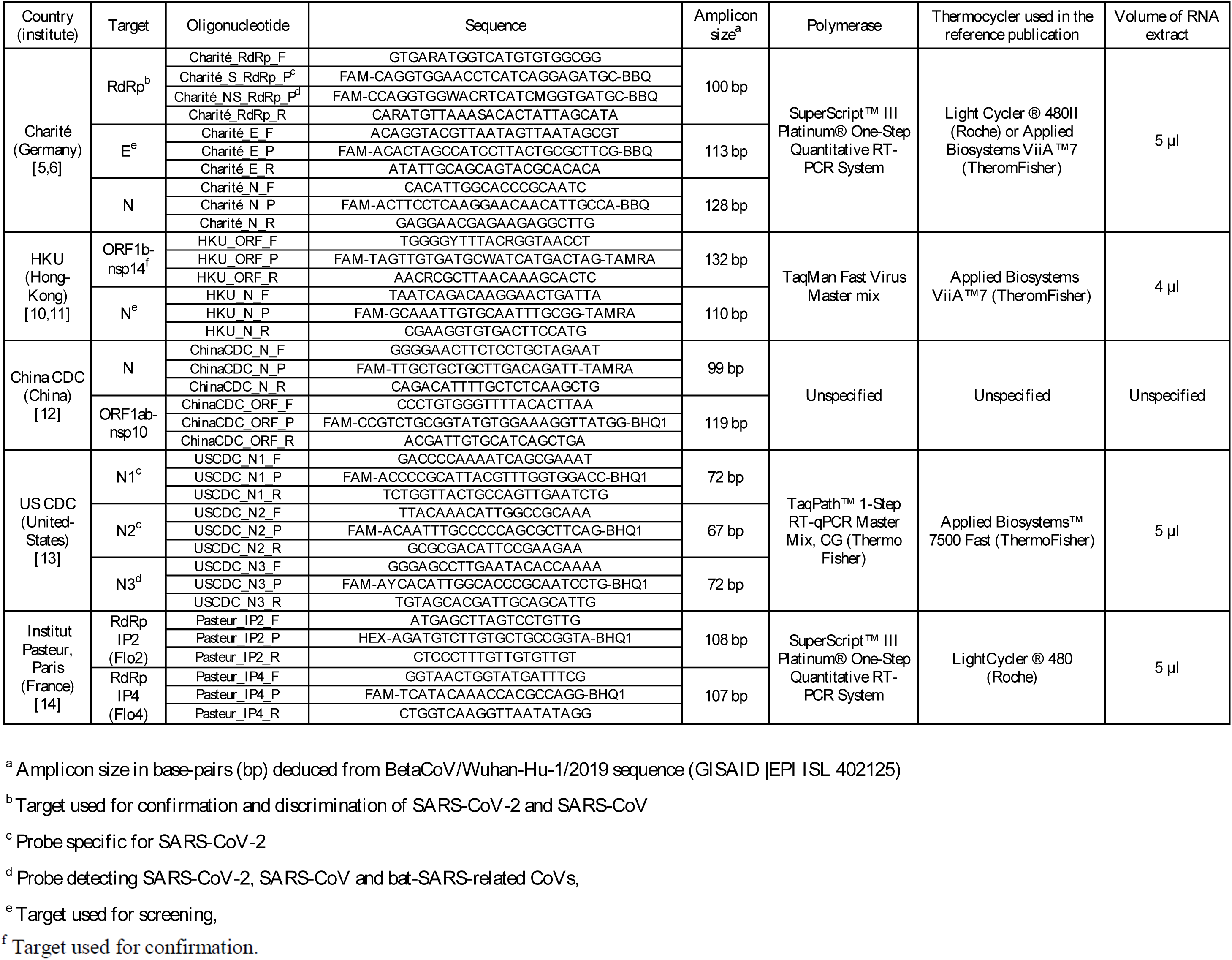
Summary of the five RT-PCR assays targeting SARS-CoV-2.

**Table 2:**
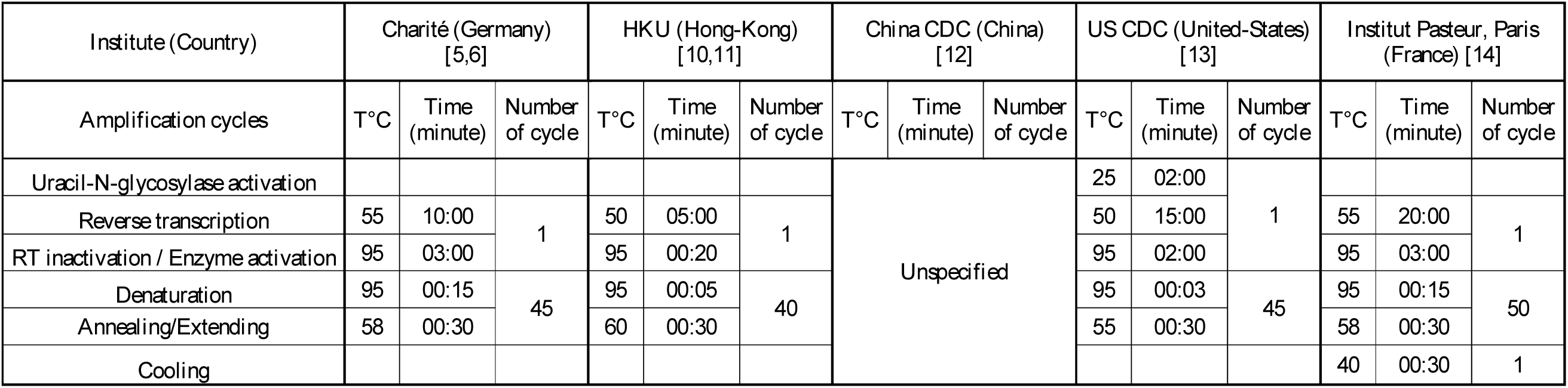
Amplification cycles of the five RT-PCR assays targeting SARS-CoV-2.

## Sensitivity comparison of the five RT-PCR assays

Sensitivity for each RT-PCR assay was first assessed using serial dilutions from 10^-3^ to 10^-9^ of SARSCoV-2 cell culture supernatants (one replicate for 10^-3^ and 10^-4^, three replicates for 10^-5^ and 10^-6^, and five replicates for 10^-7^ to 10^-9^). The E Charité and N2 US CDC assays were positive for all specimens including negative samples and negative controls (water). These false amplifications were explored (details below) but sensitivity of these assays was not further assessed.

Using specific (S) RdRp and non-specific (NS) RdRp Charité assays, all replicates of the 10^-5^ dilution (and inferior) were detected; 1/3 (S RdRp) and 3/3 (NS RdRp) of the 10^-6^ dilution replicates; and none of the 10^-7^, 10^-8^, and 10^-9^ dilutions (0/5). ORF1b and N HKU, and ORF1ab China CDC assays detected all replicates of dilutions inferior or equal to 10^-6^ and detected 4/5, 3/5 and 2/5 for 10^-7^ dilutions, respectively. None of these assays detected replicates of 10^-8^ (0/5) and 10^-9^ (0/5) dilutions. In contrast, N Charité, N China CDC, N1 and N3 US CDC, and duplex RdRp IP2/IP4 were positive for most replicates of the 10^-7^ dilutions (5/5, 5/5, 5/5, 4/5, 5/5 and 5/5, respectively) and 10^-8^ dilutions (3/5, 2/5, 4/5, 5/5, 3/5, 3/5 respectively; Figure 1). At 10^-9^ dilution, N China, N1 and N3 US CDC and duplex RdRp IP2/IP4 assays were able to detect replicates (1/5, 1/5, 2/5, 3/5, 1/5 respectively).

**Figure 1:**
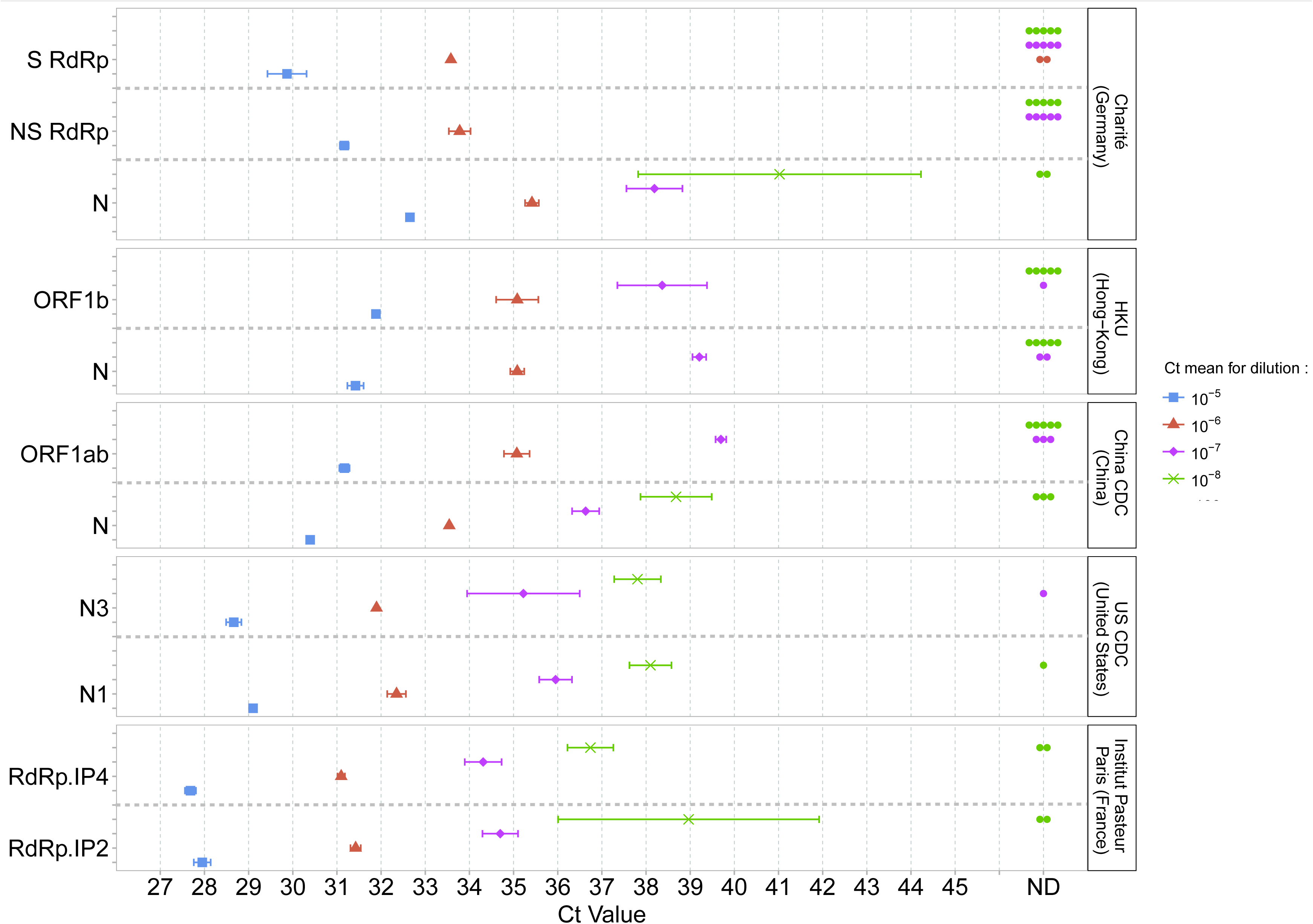
Mean Ct values and standard deviations obtained using five PCR-based methods for SARSCoV-2 detection. Serial dilutions of SARS-CoV-2 cell culture supernatants were used and are represented by a single colour (10^-5^ blue, 10^-6^ red, 10^-7^ pink, 10^-8^ green). A point in the ND (non-detected) column (Ct value axis) indicates a negative result for one replicate.

The mean cycle threshold (Ct) values obtained for each assay were then compared for dilutions 10^-5^ to 10^-8^ (Figure 1, Supplementary table 2). Since the accepted technical variability of RT-PCR is below 0.5 log_10_, we considered a difference of 2 Ct as significant. At 10^-5^ dilution, the lowest Ct value was 27.7 for RdRp IP4. No significant difference in Ct values (Ct ranged from 28.0 to 29.1) was reported with N1 and N3 US CDC, and RdRp IP2. A similar Ct profile was observed for these assays at 10^-6^ and 10^-7^ dilutions. At 10^-8^ dilution, only N Charité had significantly higher Ct values (41.0 vs 36.7 to 39.0 for N China CDC, N1 and N3 US CDC, and duplex RdRp IP2/IP4). Eight clinical samples (4 positive, 4 negative) were then tested using all RT-PCR assays to confirm the results obtained on SARS-CoV-2 cell culture supernatants. ORF1b and N HKU, ORF1ab and N China CDC, N1 and N3 US CDC, and RdRp IP2 and RdRp IP4 assays detected all 4 positive samples (Supplementary table 3). S and NS RdRp, and N Charité assays did not detect the positive sample with the lowest viral concentration. The 4 negative samples were all negative with these assays. Taken together, N China CDC, N1 and N3 US CDC as well as RdRp IP2 and IP4 were the most sensitive assays.

## Limit of detection for the most sensitive assays

The limit of detection (LoD) is defined as the lowest amount of viral genome that can be detected with a 95% hit rate. Probit analysis was applied by including five additional replicates of each dilution of the cell culture supernatants for the three most sensitive referral laboratories. Due to limited quantity of clinical samples we tested only one target of each referral laboratories: N China CDC, N1 US CDC, and RdRp IP2. LoD of N3 US CDC was not determined as this assay is not specific for SARS-CoV-2 detection and was removed from the new version of the US CDC assay [13]. We chose to determine LoD for RdRp IP2 and not IP4 as IP2 detected more replicates at the 10^-9^ dilution on cell culture supernatants.

The 95% hit rate obtained was 1.36 log_10_TCID_50_/mL [0.8; 3.09] for N China CDC, 0.44 log_10_TCID_50_/mL [0.05; 1.83] for N1 US CDC, 0.63 log_10_TCID_50_/mL [0.25; 1.9] for RdRp IP2. The differences observed were not statistically significant. For these three assays the results were confirmed by additional testing of sixteen clinical samples with low viral concentration and eight negative samples (Figure 2, supplementary table 4). N1 US CDC and RdRp IP2 had lower Ct values than N China CDC (Figure 2) but no significant differences (Ct difference <2) were observed. All negative samples were tested negative using these three assays.

**Figure 2:**
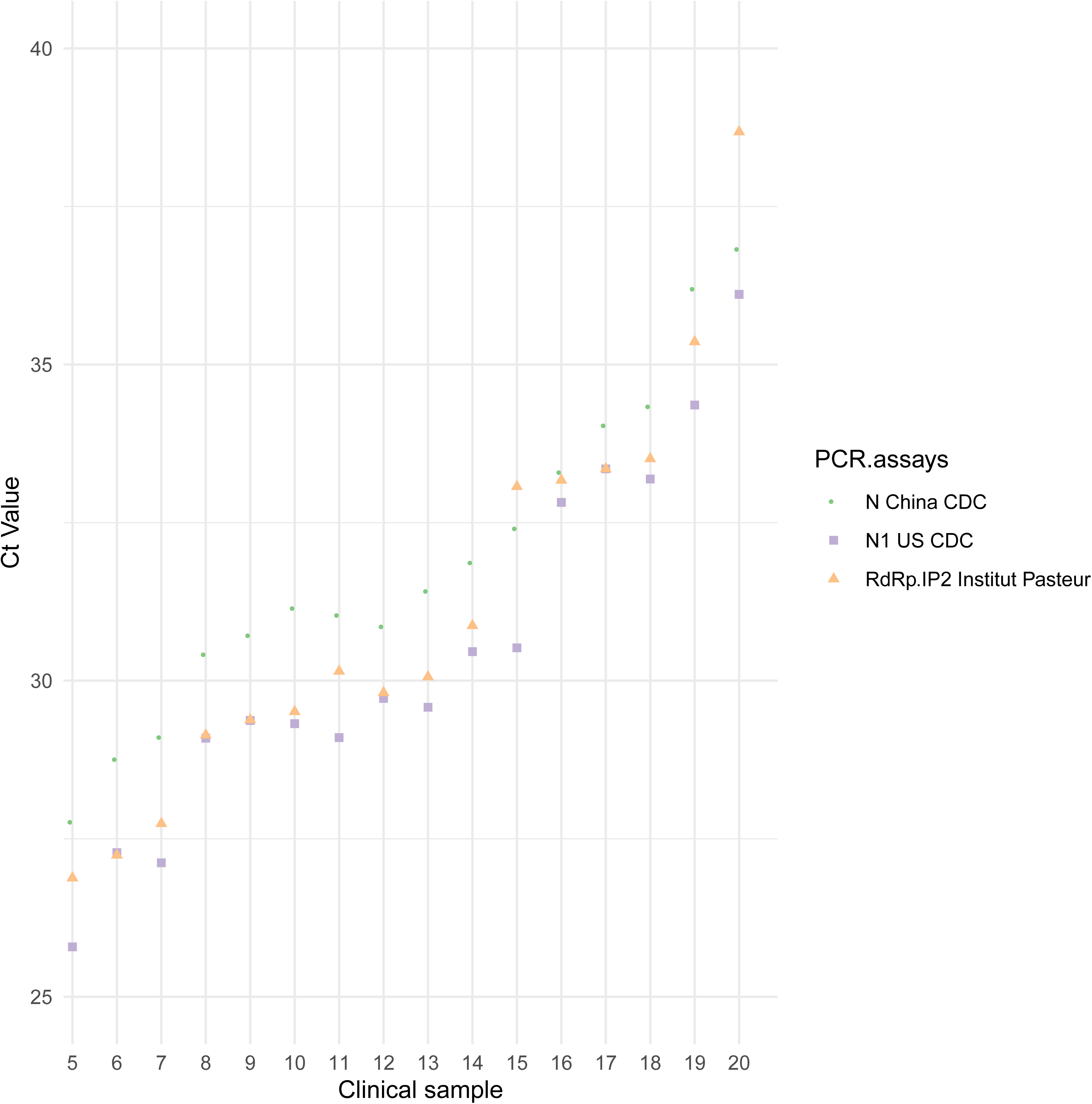
Ct values for 16 positive clinical samples using the three most sensitive assays: N China CDC (China) in green, N1 US CDC (United-States) in purple, and RdRp IP2 Institut Pasteur, Paris (France) in orange.

## Exploration of E Charité and N2 US CDC false amplifications

Since E Charité and N2 US CDC assays were positive for all specimens and replicates including negative samples and controls, we tested four additional negative clinical samples, water, and one additional clinical sample tested positive for each target. Amplicon size was analysed using Agilent DNA 1000 kit (Agilent technologies; supplementary figure 1).

For E Charité, negative samples showed two amplicons, one at 84 base pairs (bp) and one at 121 bp whereas the positive sample only had one amplicon at 121 bp, which is close to the expected size of a specific amplification (table 1). Thus, the false positive amplification obtained using E Charité might derive from a contamination (amplicon size at 121bp) but could also be associated with an aspecific amplification (amplicon size at 84bp). Using the N2 US CDC assay, negative samples showed one amplicon at 73 bp which is close to the expected size of a specific amplification (table 1). Thus, the false positive amplification obtained using N2 US CDC might be due to a contamination. Sequencing of these amplicon products should be performed for further investigation.

## Discussion

The present study is the first to compare the sensitivity of five RT-PCR-based methods developed by referral laboratories. N China CDC, N1 US CDC, and RdRp IP2 and IP4 were found to be the most sensitive assays on SARS-CoV-2 cell culture supernatants and clinical respiratory samples. Vogels et al. compared performances of SARS-CoV-2 PCR assays developed by the same referral laboratories except those from Institut Pasteur. Using RNA-spiked mock samples, they found that ORF HKU was one of the most sensitive assays [16]. Herein, ORF HKU was more sensitive than RdRp Charité but slightly less sensitive than other assays such as N1 US CDC or N HKU. Although RdRp Charité performed well for the lowest dilutions, it was nevertheless found to be less sensitive than others, a result in line with those of Vogels et al. [16]. It is worth noting that the Charité assay was the first to be published at the early stage of the pandemic [9] and has been widely used worldwide [8]. This assay was initially designed for the diagnosis of SARS-related CoVs and then optimised for SARSCoV-2 detection [5]. Thanks to this assay, an important number of COVID-19 diagnoses were made, which contributed to limit the spread of the outbreak. In line with the present results, it was reported that RdRp IP2 and RdRp IP4 sensitivity was similar when used in multiplex [14], suggesting that the Institut Pasteur assay should preferentially be used in multiplex. As previously reported [16], we identified probable primer contamination using N2 US CDC and E Charité which prevented us from further evaluation of the sensitivity. Although not observed herein, the amplification of non-specific products for ORF1 and N China CDC, and N2 and N3 US CDC has also been reported [17].

The sensitivity of other RT-PCR tests recently developed [7] should be explored in further studies. Furthermore, the specificity of each assay was not evaluated in the present study and should be determined. However, we chose to extensively assess sensitivity as a sensitive test is critical for early detection of new COVID-19 cases. The data presented herein are of prime importance to facilitate the equipment choice for all diagnostic laboratories, as well as for the development of marketed tests. Sensitive tests should be widely implemented to limit the spread of the current outbreak and prepare for the post-epidemic phase and future seasonal epidemics.

## Data Availability

All data are available upon request

